# Current infection control behaviour patterns in the UK, and how they can be improved by ‘Germ Defence’, an online behavioural intervention to reduce the spread of COVID-19 in the home

**DOI:** 10.1101/2020.06.22.20137406

**Authors:** Ben Ainsworth, Sascha Miller, James Denison-Day, Beth Stuart, Julia Groot, Cathy Rice, Jennifer Bostock, Xiao-Yang Hu, Kate Morton, Lauren Towler, Michael Moore, Merlin L Willcox, Tim Chadborn, Natalie Gold, Richard Amlôt, Paul Little, Lucy Yardley

## Abstract

**Background:** Germ Defence (https://germdefence.org/) is a freely available website providing behavioural advice for infection control within households, using behaviour change techniques. This observational study reports current infection control behaviours in the home in UK and international users of the website, and examine how they might be improved to reduce the spread of COVID-19.

**Method:** 28,285 users sought advice from four website pathways (to protect themselves generally, to protect others if the user was showing symptoms, to protect themselves if household members were showing symptoms, and to protect a household member who is at high risk) and completed outcome measures of current infection control behaviours within the home (self-isolation, social distancing, putting shopping/packages aside, wearing face-covering, cleaning and disinfecting, handwashing), and intentions to change these behaviours.

**Results:** Current user behaviours mean scores varied across all infection control measures but were between ‘sometimes’ and ‘quite often’, except handwashing (‘very often’). Behaviours were similar regardless of the website pathway used. After using Germ Defence, users recorded intentions to improve infection control behaviour across all website pathways and for all behaviours.

**Conclusions:** Self-reported infection control behaviours other than handwashing are lower than is optimal for infection prevention, although reported handwashing is much higher. The advice using behaviour change techniques in Germ Defence led to intentions to improve these behaviours. This has been shown previously to reduce the incidence, severity and transmission of infections. These findings suggest that promoting Germ Defence within national and local public health guidance could reduce COVID-19 transmission.

Section 1: What is already known on this topic

- Until a vaccine can prevent COVID-19, protective behaviours (such as social distancing, handwashing, cleaning/disinfecting) must be used to limit the spread.
- A digital behaviour change intervention to improve protective behaviours (handwashing) within the home succeeded in reducing infection transmission, healthcare utilisation and infection severity during the H1N1 pandemic (the ‘PRIMIT’ trial).
- We need to understand current levels of protective behaviour in the UK, and how to improve them, to prevent a ‘second wave’.

Section 2: What this study adds

- Our study suggests that few people are undertaking sufficient protective infection control behaviours in the home to reduce transmission
- Providing targeted digital interventions such as Germ Defence (for example through public health and primary care networks) offers a feasible method of increasing intentions to undertake these behaviours.

## Introduction

The impacts of COVID-19 must primarily be tackled through changes in behaviour undertaken by individuals and societies, until a vaccine becomes available. In many countries (including the UK), people with COVID-19 infection are instructed to remain at home, together with co-habiting family or other household members, to prevent transmission between households. This increases the risk of virus transmission within households. For example, in several environments where inter-household movement is well controlled (such as Taiwan, Ningbo and Shenzen(1–3) the virus continues to proliferate within close contacts.

To interrupt these transmission pathways, individuals must adopt ‘personal protective behaviours’(4). Such target behaviours include handwashing, disinfection of surfaces, thorough cleaning and waste disposal, social distancing within the home (where possible) and wearing situationally-appropriate personal protective equipment. A recent cohort study in Beijing, China demonstrated that performing these behaviours could dramatically reduce the likelihood of household transmission, but that the highest risk of transmission was prior to symptom onset (typically before such behaviours are performed) (5). Therefore, protective behaviours should be implemented before any household members develop symptoms. There is substantial individual variation in these behaviours, which are complex, environmentally and culturally dependent, and influenced by individual attitudes and beliefs(6). Changing such complex behaviours effectively and rapidly within the context of COVID-19 requires an approach based on behaviour change theory, evidence and extensive participatory input(7).

Specific guidance for the public on protective behaviours has been developed in many countries and is widely recommended by politicians, the media and public health/primary care networks(8). However, few behaviour change interventions have been used to support the public in these behaviours within their homes. A systematic review by our group has found evidence of only one digital intervention to date (Germ Defence) that has been shown to improve health outcomes in respiratory tract infections (RTIs) within households. Germ Defence is a mobile-friendly website that provides targeted, tailored advice about how and why users should use infection control behaviours, aiming to supplement public health guidance with evidence- and theory-based behaviour change techniques(9), optimised using extensive user feedback. In a large randomised controlled trial of 20066 people (the PRIMIT trial) during the previous H1N1 pandemic (10), those randomised to use Germ Defence had reduced frequency and severity of respiratory infections and reduced transmission to household members.

Germ Defence was rapidly adapted for the COVID-19 pandemic by a team of medical, public health and behaviour change experts, and public contributors. It was then disseminated through multiple pathways (primarily but not exclusively in the UK) including public health and primary care networks (for example, by texting the website link to patients via GP practices), national and local press, television coverage and social media.

In this study, we aimed to:

1. Examine current infection control behaviours (recorded in 28,285 people who used Germ Defence between the 6^th^ May and 24^th^ May 2020).
2. Examine whether using Germ Defence improves intentions to change behaviour to control infection transmission.

## Method

### Design

This was a pragmatic, cross-sectional observational study of anonymous participant data from an active behavioural intervention. Consent was assumed from website usage and acknowledged in the website privacy policy. The study received ethical approval from University of Bath (PREC reference 20-088).

### Participants and Data

The data analysed were collected from users of the Germ Defence website between May 6th and May 24th 2020 (selected period during which the Germ Defence website was disseminated in the UK and underwent no substantial modification). During this period 70,566 website hits were recorded, with 53,125 users completing the introductory content (first three pages) and 28,285 people completing the core module, which included measures of current and intended behaviour. Website usage and engagement data was collected using Google Analytics embedded in the site. A full consort diagram of usage is presented in Figure 1.

**Figure 1.**
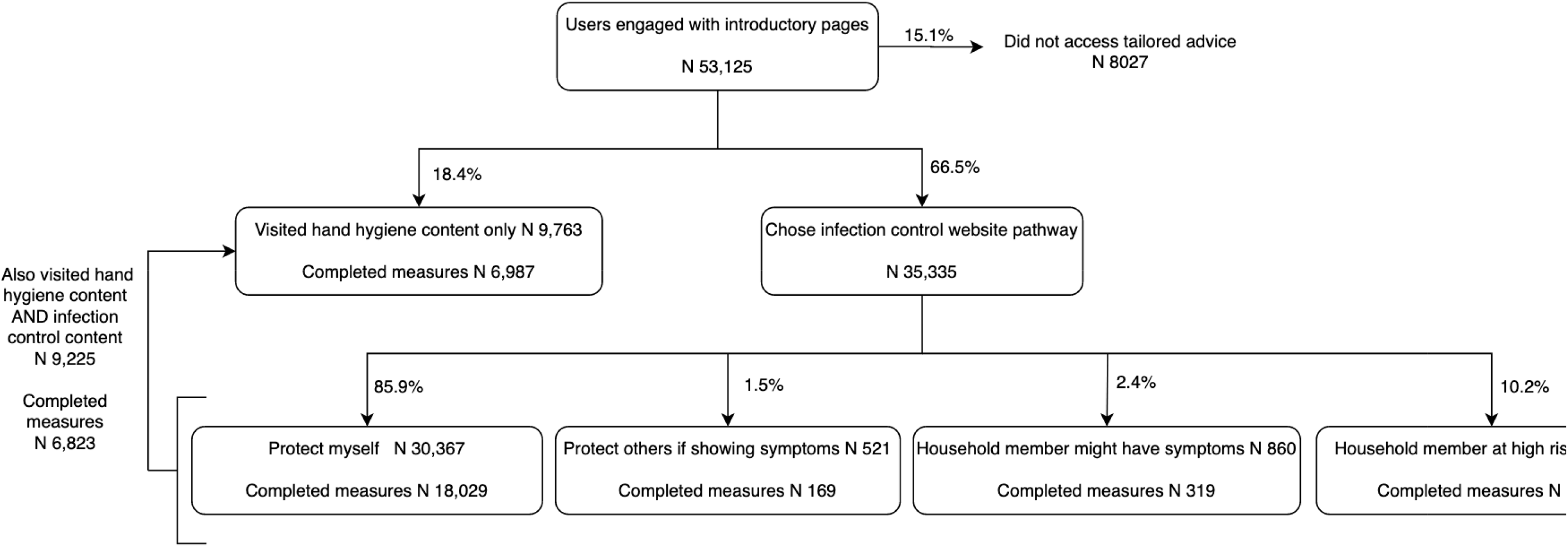
Flow diagram of the number of users who visited each part of Germ Defence.

Behavioural measures were recorded through self-report questions within the website. Measures were all scored on a Likert scale with answers of 1 (Almost never), 2 (sometimes), 3 (quite often), 4 (very often) and 5 (almost always). Users could also answer ‘Not Applicable’ (for example, if they lived alone and therefore did not need to socially isolate within their household). Measures were recorded for current behaviour and intended behaviour.

### Infection control measures

#### Social distancing

When you were/are with them, how often were you/do you plan to be more than 2 metres/6 feet away from the people you live with?

#### Cleaning/Disinfecting

How often did you/do you plan to clean things that might have viruses on them?

#### Putting shopping/packages aside

How often did you/do you plan to put something aside for at least 1 day that might have viruses on it?

#### Self-isolating

How often did you/do you plan to spend time in a room on your own?

#### Wearing face coverings

How often did you/do you plan to wear a face covering and glasses (and safely remove and clean them) when you are in the same room as other people?

### Detailed handwashing measures

How often did you/do you plan to wash your hands…:

#### Before snacking

Before I ate/eat with my fingers e.g. snack, fruit or sweets?

#### After coming home

When I came/come into a house e.g. after work, shopping, travelling?

#### After coughing

After blowing my nose or sneezing/coughing on my hands?

#### After coming into contact with possible carrier

After I had been/being close to someone who may have a virus (within 6 feet)

#### After touching something

After touching something that lots of other people have touched e.g. doors, money or handrails?

### Helpfulness of site

How strongly do you agree or disagree that Germ Defence was helpful to you? (1-10)

### Intervention

Germ Defence content was developed using theoretical modelling and qualitative research(11), in line with the person based approach(12), drawing principally on the theory of planned behaviour(13), Leventhal’s common-sense model of illness(14) and protection motivation theory(15). Intervention content, design and structure were optimised iteratively using in-depth qualitative think-aloud interviews with public contributors (JB,CR) and members of the general public in order to ensure the intervention was accessible, credible and motivating for as many people as possible(12).

Based on process evaluations of the original randomised controlled trial(10) and a previous public dissemination(16), Germ Defence has been updated and streamlined for use during the COVID-19 outbreak, including broadening the infection control behaviours that were recommended. The intervention is a single session, designed to be easily accessible with no sign-up or password required. A structured outline of content is available in Table 1. The consent process was placed within the website privacy policy. Data collection was unobtrusive and kept to a minimum to reduce dropout.

**Table 1.** A detailed outline of Germ Defence content and structure

| Germ Defence Component | Component Details |
| --- | --- |
| **Introductory content**<br>(3 pages) | Introductory pages seek to increase users' perceived risk by emphasising the personal and social health consequences of contracting COVID-19. These are followed by messages to increase skills and confidence to reduce exposure to the virus. |
| **Website pathway selection**<br>(2 pages) | To allow users to choose the advice they consider most personally relevant, the intervention is structured so that users initially select between two components of interest: Handwashing and Reducing Illness. The 'Reducing Illness' component is again tailored such that a user selects one of four streams of content (each lasting 11 pages) that is relevant to the user's situation: 1: to protect themselves generally; 2: to protect others if the user was showing symptoms; 3: to protect themselves if household member(s) showed symptoms; or 4: to protect a household member who is at high risk. The advice is tailored in this way to encourage users to adopt behaviours appropriate to the perceived level and pattern of risk in their household. For example, users in the 'protect themselves generally' would vary from very low to very high risk. It was not possible to provide specific tailored advice for every household combination of risks and resources (for example, based on the need and potential for household members to self-isolate within the home); therefore, Germ Defence aimed to educate users to adopt behaviours that were appropriate and feasible for their own circumstances. |
| **Tailored infection control behaviour advice**<br>(7 pages) | Clear and detailed advice is then provided for self-isolating, social distancing, disinfecting/cleaning, wearing face-coverings, and putting items aside that may have viruses on them such as shopping/packages. Advice is provided to the extent that users feel is appropriate for the perceived risk. These pages also contain ideas and information on how to structure the home and engage in behaviours safely. The handwashing component provides advice focused on handwashing that is relevant to all groups over 5 pages. |
| **Goal setting advice** (3 pages) | Both the Handwashing and Reducing Illness components contain goal-setting sections where users indicate their behaviour over the past week, view a motivational message, and then plan their behaviour for the future. Users who do not select any improvement are encouraged to review their plan. After completing either Handwashing or Reducing Illness components, users are asked how helpful they found the website. |
| **Additional information** | Users are then able to revisit the first two components, choose from two additional components with more detailed information about the same behaviours (e.g., how to social distance with young children, how to stop touching your face), or view details about the website. |

**Note:** The website, and all associated content, can be accessed for free: https://germdefence.org/

### Statistical analysis

We included data from all users who accessed the website between May 6th and May 24th 2020. Website usage data was used to explore user locations, time spent on the website, website return rate and content visited.

For analysis, users were grouped according to the tailored website pathway they selected within the ‘Reducing illness’ component (‘Protect myself generally’ vs. ‘Protect others if I am showing symptoms’ vs ‘Protect myself if a household member has symptoms’ vs. ‘Protect a household member at high risk’). Users could also view the Handwashing component which was relevant to all groups. If they did not view ‘Reducing illness’ they were not included in group comparisons, but handwashing responses were still recorded. Users could complete more than one type of tailored pathway but we only analysed responses for the pathway that was selected first.

Behavioural measures were analysed individually, and also collapsed together to form an ‘average infection control behaviour’ score. When users completed a plan more than once (for example, if they received website feedback that their initial plan could be further improved) the ‘final’ plan was used.

If users did not think a behaviour was relevant to them (for example, they lived alone so did not need to socially isolate, or were not able to socially isolate from young children) they could answer ‘not applicable’, which was coded as missing data and not included in analysis.

Linear regression compared between-group scores for behaviour. Models comparing between-group scores for intentions controlled for current behaviour. Paired t-tests were used to examine the difference between current behaviour and intended behaviour within groups.

## Results

### Usage of the Germ Defence website

We considered data from 53,125 users who completed at least the initial introductory website pages. Users accessed Germ Defence from 129 countries (a full consort diagram of usage is presented in Figure 1). 83.7% of users were from the UK (England 75.6%, Scotland 4.2%, Wales 2.8%, Northern Ireland 1.1%, other UK 0.2%). Mean usage lasted 8 minutes 28 seconds on the intervention, and mean number of pages viewed was 19.9. 54.1% of recorded sessions lasted longer than one minute. 54.0% of users accessed Germ Defence using a mobile device, 31.0% with a tablet and 15.0% with a desktop or laptop computer. 10.6% of users were ‘return users’ visiting for a second time. Detailed usage for each website component is presented in Figure 1.

Overall mean helpfulness of the website was rated as 7.77 out of 10 (SD 2.31).

### Infection control behaviours and intended behaviours in users of Germ Defence

All groups (protect themselves generally, protect others if the user was showing symptoms, protect themselves if household members were showing symptoms, and protect a household member who is high risk) reported using most current infection behaviours sometimes/quite often within the home. Overall, users reported they would wear a face covering almost never/sometimes (M 1.61, SD 1.19) and would socially distance sometimes/quite often (M 2.40, SD 1.22). Users reported socially isolating in their own room sometimes/quite often (M 2.78, SD 1.29) and putting packages/shopping aside sometimes/quite often (M 2.75, SD 1.55). Users reported cleaning/disinfecting quite often/very often (M 3.17, SD 1.18).

Frequency of the five infection control behaviours from the ‘Reducing Illness’ pathway within each group is reported in Table 2 (with handwashing reported separately below), as well as mean differences and confidence intervals of group comparisons (each group vs. ‘Protect themselves generally’ group). The frequency of behaviours did not vary appreciably between groups; numerically, the ‘Protect themselves generally’ group were least likely to socially distance (M 2.39, SD 1.22). People in the ‘Protect others if user showing symptoms’ group were least likely to clean/disinfect (M 2.95, SD 1.26) and put aside shopping/packages (M 2.39, SD 1.48) but most likely to wear a face covering (M 1.91, SD 1.36). People in ‘Protect themselves if household members showing symptoms’ were most likely to maintain social distance (M 2.57, SD 1.23), and users in ‘Protect household members at high risk’ were least likely to stay in their own room (M 2.64, SD 1.16) and least likely to wear a face covering (M 1.42, SD 0.99).

**Table 2.**
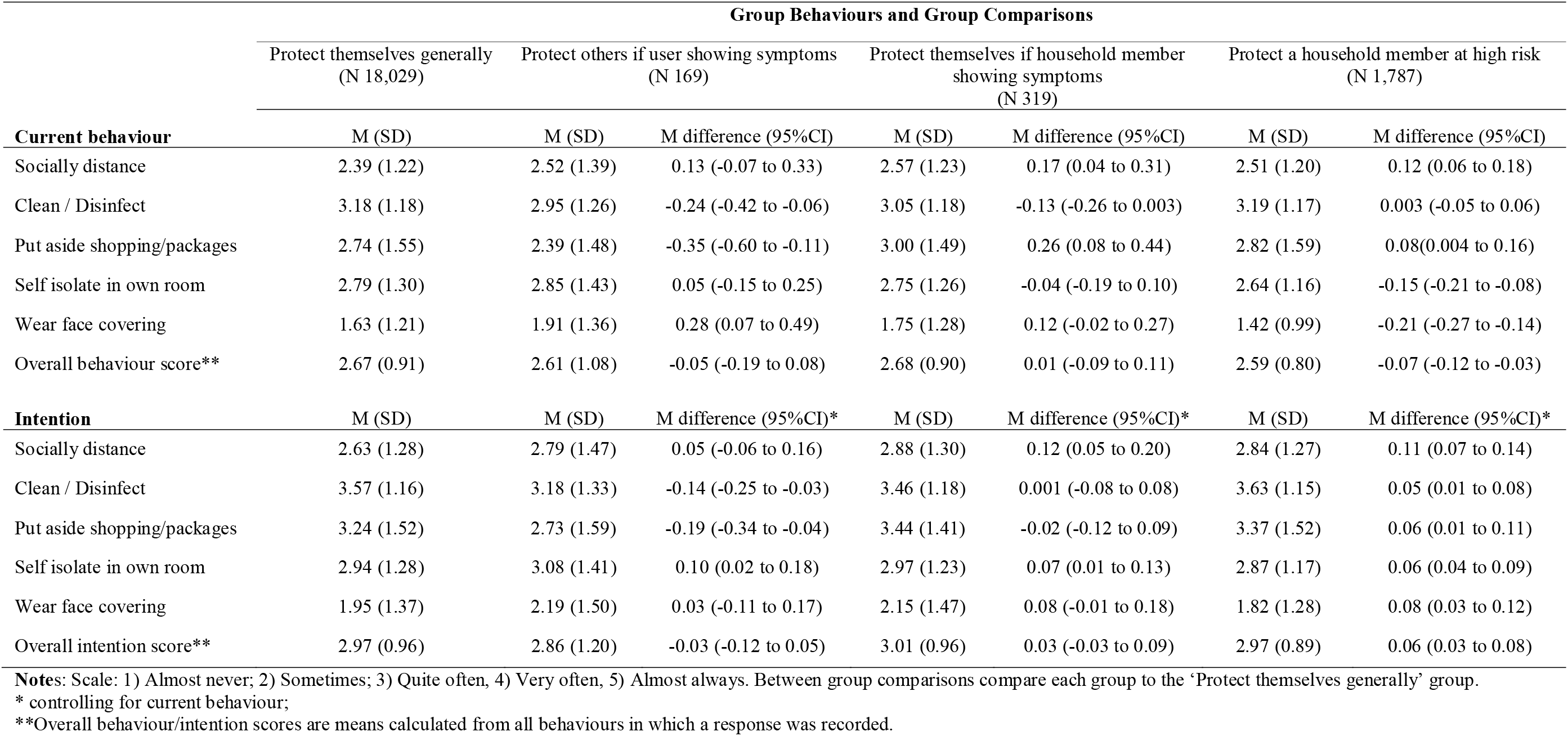
Current and intended infection control behaviours.

| Group Behaviours and Group Comparisons |  |  |  |  |  |  |  |
| --- | --- | --- | --- | --- | --- | --- | --- |
|  | Protect themselves generally<br>(N 18,029) | Protect others if user showing symptoms<br>(N 169) |  | Protect themselves if household member<br>showing symptoms<br>(N 319) |  | Protect a household member at high risk<br>(N 1,787) |  |
| **Current behaviour** | M (SD) | M (SD) | M difference (95%CI) | M (SD) | M difference (95%CI) | M (SD) | M difference (95%CI) |
| Socially distance | 2.39 (1.22) | 2.52 (1.39) | 0.13 (-0.07 to 0.33) | 2.57 (1.23) | 0.17 (0.04 to 0.31) | 2.51 (1.20) | 0.12 (0.06 to 0.18) |
| Clean / Disinfect | 3.18 (1.18) | 2.95 (1.26) | -0.24 (-0.42 to -0.06) | 3.05 (1.18) | -0.13 (-0.26 to 0.003) | 3.19 (1.17) | 0.003 (-0.05 to 0.06) |
| Put aside shopping/packages | 2.74 (1.55) | 2.39 (1.48) | -0.35 (-0.60 to -0.11) | 3.00 (1.49) | 0.26 (0.08 to 0.44) | 2.82 (1.59) | 0.08(0.004 to 0.16) |
| Self isolate in own room | 2.79 (1.30) | 2.85 (1.43) | 0.05 (-0.15 to 0.25) | 2.75 (1.26) | -0.04 (-0.19 to 0.10) | 2.64 (1.16) | -0.15 (-0.21 to -0.08) |
| Wear face covering | 1.63 (1.21) | 1.91 (1.36) | 0.28 (0.07 to 0.49) | 1.75 (1.28) | 0.12 (-0.02 to 0.27) | 1.42 (0.99) | -0.21 (-0.27 to -0.14) |
| Overall behaviour score** | 2.67 (0.91) | 2.61 (1.08) | -0.05 (-0.19 to 0.08) | 2.68 (0.90) | 0.01 (-0.09 to 0.11) | 2.59 (0.80) | -0.07 (-0.12 to -0.03) |
| **Intention** | M (SD) | M (SD) | M difference (95%CI)* | M (SD) | M difference (95%CI)* | M (SD) | M difference (95%CI)* |
| Socially distance | 2.63 (1.28) | 2.79 (1.47) | 0.05 (-0.06 to 0.16) | 2.88 (1.30) | 0.12 (0.05 to 0.20) | 2.84 (1.27) | 0.11 (0.07 to 0.14) |
| Clean / Disinfect | 3.57 (1.16) | 3.18 (1.33) | -0.14 (-0.25 to -0.03) | 3.46 (1.18) | 0.001 (-0.08 to 0.08) | 3.63 (1.15) | 0.05 (0.01 to 0.08) |
| Put aside shopping/packages | 3.24 (1.52) | 2.73 (1.59) | -0.19 (-0.34 to -0.04) | 3.44 (1.41) | -0.02 (-0.12 to 0.09) | 3.37 (1.52) | 0.06 (0.01 to 0.11) |
| Self isolate in own room | 2.94 (1.28) | 3.08 (1.41) | 0.10 (0.02 to 0.18) | 2.97 (1.23) | 0.07 (0.01 to 0.13) | 2.87 (1.17) | 0.06 (0.04 to 0.09) |
| Wear face covering | 1.95 (1.37) | 2.19 (1.50) | 0.03 (-0.11 to 0.17) | 2.15 (1.47) | 0.08 (-0.01 to 0.18) | 1.82 (1.28) | 0.08 (0.03 to 0.12) |
| Overall intention score** | 2.97 (0.96) | 2.86 (1.20) | -0.03 (-0.12 to 0.05) | 3.01 (0.96) | 0.03 (-0.03 to 0.09) | 2.97 (0.89) | 0.06 (0.03 to 0.08) |

**Notes:** Scale: 1) Almost never; 2) Sometimes; 3) Quite often, 4) Very often, 5) Almost always. Between group comparisons compare each group to the 'Protect themselves generally' group.
\* controlling for current behaviour;
\*\*Overall behaviour/intention scores are means calculated from all behaviours in which a response was recorded.

Table 2 shows some small differences in how often participants planned to perform behaviours in the future (corrected for levels of current behaviour) between groups. Compared to people in the ‘Protect themselves generally’ group, people showing symptoms planned to clean/disinfect and put aside less frequently, but planned to self-isolate more frequently. People in the ‘Protect themselves from household member with symptoms’ group planned to socially distance and self-isolate more frequently than those in the ‘Protect themselves generally’ group. People looking to protect a high-risk household member planned to conduct all of the behaviours slightly more frequently than the ‘Protect themselves generally’ group.

Paired t-test comparisons examined differences between current and planned behaviours after using the Germ Defence website. Mean difference scores for each group and 95% confidence intervals are reported in Table 3. The difference between intended and current behaviour was largest for cleaning/disinfecting (M 0.38, 0.37 to 0.39) and putting aside shopping/packages (M 0.49, 0.47 to 0.50), and lowest for self-isolating (M 0.15, 0.14 to 0.16). Overall, infection control behaviours increased (M 0.30, 0.29 to 0.31).

**Table 3.** Group differences between behaviour and intention.

|  | Protect themselves generally<br>(N 17,239) | Protect others if user<br>showing symptoms<br>(N 157) | Protect themselves if household<br>member showing symptoms<br>(N 303) | Protect a household<br>member at high risk<br>(N 1,693) | Overall<br>(N 19,392) |
| --- | --- | --- | --- | --- | --- |
| Socially distance | 0.22 (0.21 to 0.23) | 0.26 (0.11 to 0.40) | 0.33 (0.24 to 0.42) | 0.31 (0.28 to 0.35) | 0.23 (0.22 to 0.24) |
| Clean / Disinfect | 0.38 (0.37 to 0.39) | 0.30 (0.17 to 0.44) | 0.41 (0.31 to 0.51) | 0.43 (0.39 to 0.47) | 0.38 (0.37 to 0.40) |
| Put aside shopping/packages | 0.49 (0.47 to 0.50) | 0.39 (0.24 to 0.54) | 0.41 (0.31 to 0.51) | 0.53 (0.48 to 0.58) | 0.49 (0.47 to 0.50) |
| Self isolate in own room | 0.14 (0.13 to 0.15) | 0.23 (0.11 to 0.36) | 0.21 (0.14 to 0.29) | 0.22 (0.19 to 0.25) | 0.15 (0.14 to 0.16) |
| Wear face covering | 0.28 (0.27 to 0.30) | 0.29 (0.12 to 0.47) | 0.35 (0.25 to 0.46) | 0.37 (0.33 to 0.42) | 0.29 (0.28 to 0.29) |
| Average infection control score | 0.29 (0.29 to 0.30) | 0.27 (0.16 to 0.38) | 0.32 (0.25 to 0.40) | 0.36 (0.33 to 0.39) | 0.30 (0.29 to 0.31) |

**Note:** Group Ns are taken across all behaviours.

Handwashing behaviour is reported in Table 4. Mean current handwashing behaviour was higher than other infection control behaviours (M 4.04, SD 0.84) with reported intended behaviour consistently higher (Mean increase 0.41, 95%CI 0.40 to 0.42).

**Table 4:**
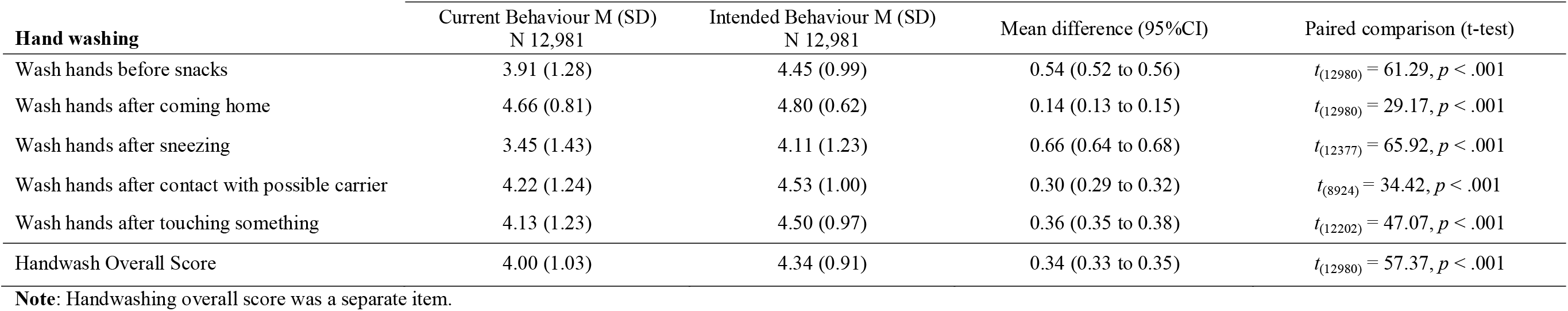
Paired comparisons between current and intended handwashing behaviour.

| **Hand washing** | Current Behaviour M (SD)<br>N 12,981 | Intended Behaviour M (SD)<br>N 12,981 | Mean difference (95%CI) | Paired comparison (t-test) |
| --- | --- | --- | --- | --- |
| Wash hands before snacks | 3.91 (1.28) | 4.45 (0.99) | 0.54 (0.52 to 0.56) | $t_{(12980)} = 61.29, p < .001$ |
| Wash hands after coming home | 4.66 (0.81) | 4.80 (0.62) | 0.14 (0.13 to 0.15) | $t_{(12980)} = 29.17, p < .001$ |
| Wash hands after sneezing | 3.45 (1.43) | 4.11 (1.23) | 0.66 (0.64 to 0.68) | $t_{(12377)} = 65.92, p < .001$ |
| Wash hands after contact with possible carrier | 4.22 (1.24) | 4.53 (1.00) | 0.30 (0.29 to 0.32) | $t_{(8924)} = 34.42, p < .001$ |
| Wash hands after touching something | 4.13 (1.23) | 4.50 (0.97) | 0.36 (0.35 to 0.38) | $t_{(12202)} = 47.07, p < .001$ |
| Handwash Overall Score | 4.00 (1.03) | 4.34 (0.91) | 0.34 (0.33 to 0.35) | $t_{(12980)} = 57.37, p < .001$ |

**Note:** Handwashing overall score was a separate item.

## Discussion

### Summary of findings

Germ Defence was accessed by a large number of users across 129 countries, primarily from the UK. This demonstrates substantial public interest in adopting appropriate infection control behaviours in the home during the COVID-19 pandemic. After using Germ Defence, all groups reported intentions to increase the frequency of their infection control behaviours, including handwashing.

Except for handwashing, self-reported infection control behaviours in the home were only reported ‘sometimes/quite often’ regardless of whether people were seeking to protect themselves, concerned about demonstrating COVID-19 symptoms, had a household member showing symptoms, or were seeking to protect a high-risk household member. The frequency of wearing face coverings was consistently lowest of the behaviours, while cleaning/disinfecting was the most frequently reported of the behaviours outside of handwashing. All of these infection control behaviours were reported to be performed much less frequently than was handwashing.

As would be expected, certain behaviours and intentions varied according to the circumstances of groups – for example, people seeking to protect a high risk household member reported the intention to perform all behaviours slightly more frequently, while people seeking to protect themselves from a household member with symptoms were more likely to self-isolate in their own room.

### Implications of the study

This study provides the first up-to-date analysis of infection control behaviours and intentions across the UK, in a large sample during the COVID-19 pandemic. Within-household transmission is likely to be an increasingly important determinant of morbidity and mortality as infection control measures become established in external, public environments(6,17). Therefore, understanding current infection control behaviours and to what extent they can be improved within homes is vital to continue to control the pandemic.

Self-reported infection control behaviours other than handwashing are lower than is optimal for infection prevention; even in Germ Defence users who were likely more motivated and more likely to engage in protective behaviours than the general population (as they were seeking additional information)(18). Increasing engagement in these behaviours is important as societal restrictions are released and perceived risk reduces(19).

A concerted effort to improve household infection control behaviours across the UK population is likely to be an efficient use of health resource, both to reduce current rates of infection and to prevent the likelihood and severity of future outbreaks (a ‘second wave’). Handwashing behaviours are already relatively high – perhaps due to existing familiarity with the behaviour, supported by a focus in public health advice on increasing handwashing in earlier stages of the pandemic. Therefore, other infection control behaviours must be targeted by theory and evidence-based interventions such as Germ Defence.

Germ Defence users reported intentions to increase the frequency of infection control behaviours over their current rates. Although such intentions potentially misrepresent the observed behavioural change after an intervention (the ‘intention-behaviour gap’(20)) there is good evidence that Germ Defence overcomes this. Analysis of comparable data from the PRIMIT trial handwashing intervention showed slightly smaller behaviour/intention differences (effect size d_z_ 0.45). This change was sufficient to cause reduced infection transmission and severity within households after 16 weeks(10). Comparable data during the current pandemic (Reducing Illness behaviours d_z_ 0.53; handwashing d_z_ 0.50) shows a slightly larger effect across a broader range of behaviours that may have a larger impact on infection rates.

Given the current rates of infection control behaviours within the home, even within a motivated sample (including symptomatic participants, or with household members at high risk), it is vital to address barriers to engaging in these behaviours. For example, people living in crowded, working households are more likely to come into contact with the virus(5), and may also find it difficult to self-isolate. Similarly, cultural differences within households, financial challenges, or caring responsibilities may cause barriers to social distancing(6). This may be why the improvement between behaviours and intentions was smaller for social distancing and self-isolation compared to behaviours less likely to have significant barriers to overcome (cleaning/disinfecting, and putting packages aside). Research should explore how to support these behaviours for as many households as possible.

### Limitations of the study

As a cross-sectional observation of an active intervention, Germ Defence lacks longitudinal follow-up. Care must be taken when interpreting findings within the rapidly changing context of the COVID-19 pandemic. Although our data suggests that the frequency of infection control behaviours did not change during the study (see Appendix 1), we do not have comparable data from before the pandemic. Our method of categorisation using website pathways may not be accurate for some users, or might overlook individual differences (for example, some users in the ‘Protect myself’ category may be extremely high risk).

Our data may not be a representative sample from the wider UK population, for several reasons. Firstly, users of Germ Defence are likely to be more motivated and report higher frequencies of infection control behaviours. Secondly, although analytic data indicates that the large majority of users of the intervention were from the UK, we could not identify non-UK users within behavioural data. Finally, self-reported infection control behaviours may not be accurate reflections of actual behaviours occurring within households.

However, none of these limitations affect our main findings; indeed, people are prone to over-report protective behaviours, further highlighting the need for improvement.

## Conclusions

Our findings show substantial room for improvement in protective behaviours across the UK, even in our motivated, self-selected sample, as societal restrictions are eased. People are not sufficiently self-isolating within the home in order to prevent household transmission, even when a household member or they themselves are demonstrating COVID-19 symptoms. Our finding that Germ Defence improved intentions to undertake protective behaviours suggests that promoting evidence-based behaviour change interventions might improve these behaviours and reduce both transmission within households and the incidence and severity of infections.

Germ Defence (https://germdefence.org) is a scalable, evidence-based, acceptable and free public health intervention with negligible safety risk, which could be included in public heath guidance and promoted via primary care networks at minimal cost for wide population coverage. This would add to the limited support provided to the public to adhere to these important behaviours and mitigate their personal risk, and complement other population health protection interventions.

## Data Availability

All time-stamped data files used in analysis (and analysis scripts) will be made available upon final manuscript publication.

## Acknowledgements

We thank all who have supported and disseminated Germ Defence, including the University of Bath (Andy Dunne) and NIHR Bristol Health Protection Research Unit (Helen Bolton and Clare Thomas). We thank all who have assisted in the translation of Germ Defence into other languages (documented here: http://lang.germdefence.org/). We also thank the many citizen scientists and public contributors who assisted in the development of the Germ Defence intervention.

## Competing Interests

All authors have completed the *Unified Competing Interest form* (available on request from the BA) and declare: no support from any organisation for the submitted; no financial relationships with any organisations that might have an interest in the submitted work in the previous three years, no other relationships or activities that could appear to have influenced the submitted work.

## Contributors

Conceived the study: BA, SM, LY.

Study Design: BA, LY.

Analysed the data: BA, BS, JG.

Interpreted the data: All authors

Developed the intervention: All authors

Drafted the manuscript: BA

Reviewed the manuscript and approved the content: All authors

Met authorship criteria: All authors.

## Funding

The study was funded by the UKRI/MRC Rapid Response Call: UKRI CV220-009.

The Germ Defence intervention was hosted by the Lifeguide Team, supported by the NIHR Biomedical Research Centre, University of Southampton. LY is a National Institute for Health Research (NIHR) Senior Investigator and theme lead for University of Southampton Biomedical Research Centre. LY and RA are affiliated to the National Institute for Health Research Health Protection Research Unit (NIHR HPRU) in Behavioural Science and Evaluation of Interventions at the University of Bristol in partnership with Public Health England (PHE). MLW is a NIHR Academic Clinical Lecturer, under grant CL-2016-26-005. The views expressed are those of the author(s) and not necessarily those of the NHS, the NIHR, the Department of Health or Public Health England. The funders had no role in the design of the study, collection, analysis, and interpretation of data or in writing the manuscript.

## Ethics and Data-sharing statement

Consent was assumed from website usage and acknowledged in the website privacy policy. The study received ethical approval from University of Bath (PREC reference 20-088). All time-stamped data files used in analysis (and analysis scripts) will be made available upon final.

## Transparency Declaration

The lead authors (BA and LY) confirm that the manuscript is an honest, accurate, and transparent account of the study being reported; that no important aspects of the study have been omitted; and any discrepancies have been explained.

## Exclusive License

The Corresponding Author has the right to grant on behalf of all authors and does grant on behalf of all authors, a worldwide licence to the Publishers and its licensees in perpetuity, in all forms, formats and media (whether known now or created in the future), to i) publish, reproduce, distribute, display and store the Contribution, ii) translate the Contribution into other languages, create adaptations, reprints, include within collections and create summaries, extracts and/or, abstracts of the Contribution and convert or allow conversion into any format including without limitation audio, iii) create any other derivative work(s) based in whole or part on the on the Contribution, iv) to exploit all subsidiary rights to exploit all subsidiary rights that currently exist or as may exist in the future in the Contribution, v) the inclusion of electronic links from the Contribution to third party material where-ever it may be located; and, vi) licence any third party to do any or all of the above. All research articles will be made available on an open access basis (with authors being asked to pay an open access fee—see http://www.bmj.com/about-bmj/resources-authors/forms-policies-and-checklists/copyright-open-access-and-permission-reuse). The terms of such open access shall be governed by a Creative Commons licence— details as to which Creative Commons licence will apply to the research article are set out in our worldwide licence referred to above.

## Dissemination

It is not possible to disseminate these results to study participants as they are anonymous.

## Appendix 1: Mean infection control behaviour frequency (not including handwashing) during the period of the study

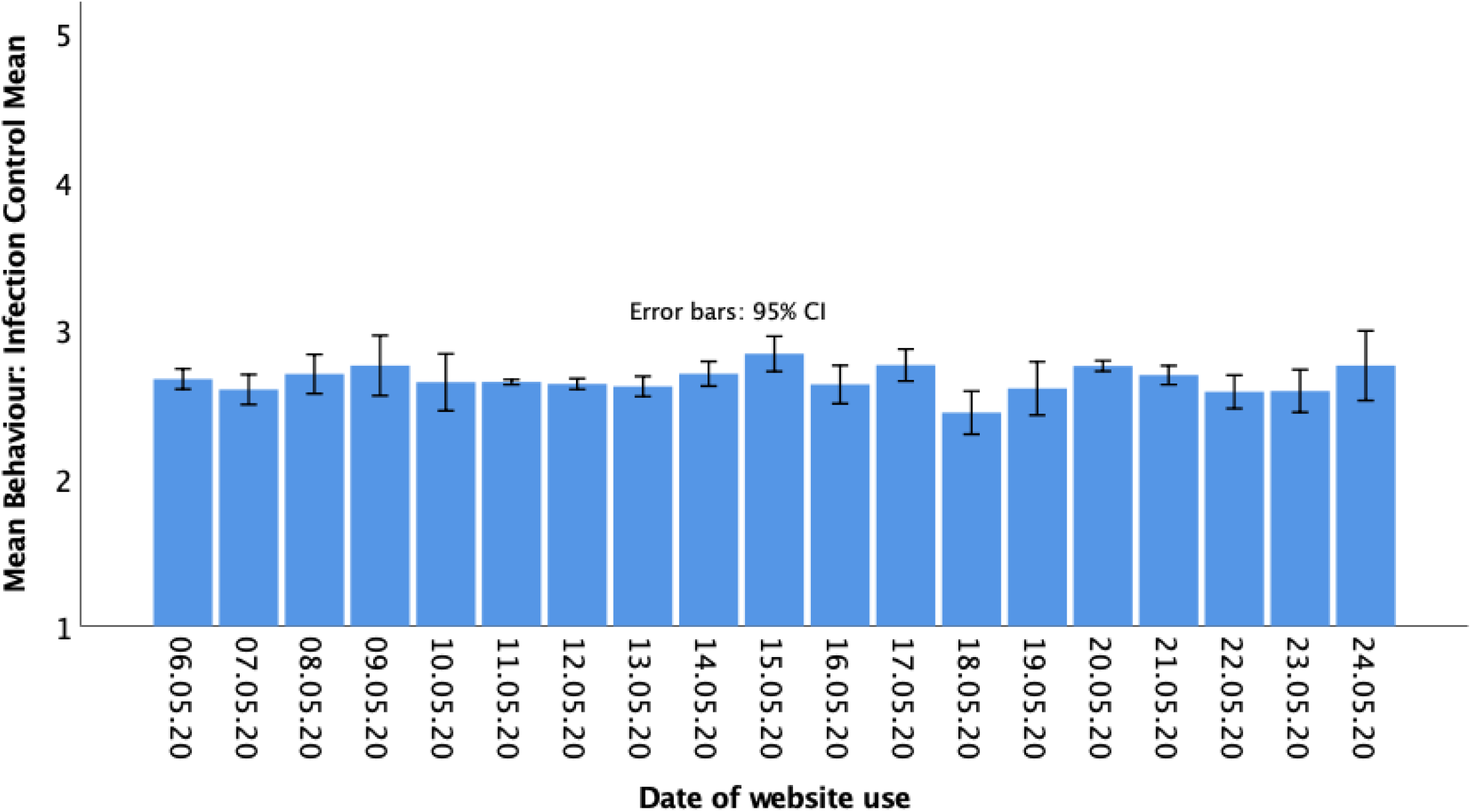

## Notes

### Competing Interest Statement

The authors have declared no competing interest.

### Author Declarations

The study received ethical approval from University of Bath (PREC reference 20-088).

